# Pitfalls in 2HG detection with TE-optimized MRS at 3T

**DOI:** 10.1101/2025.03.31.25324828

**Authors:** Seyma Alcicek, Dunja Simicic, Lindsay Blair, Max Saint-Germain, Helge J. Zöllner, Christopher W. Davies-Jenkins, Matthias Holdhoff, John Laterra, Chetan Bettegowda, Karisa C. Schreck, Doris D. Lin, Peter B. Barker, David O. Kamson, Georg Oeltzschner

## Abstract

**Background and Purpose:** In-vivo magnetic resonance spectroscopy (MRS) of 2-hydroxyglutarate (2HG) may provide diagnostic and monitoring biomarkers in isocitrate dehydrogenase (IDH)-mutated glioma. A previous meta-analysis has shown good diagnostic accuracy of TE-optimized PRESS for IDH-mutated glioma, but most studies feature IDH-wildtype glioma as a comparison. However, when considering newly identified brain lesions that may mimic glioma, full characterization of its diagnostic utility should also consider the accuracy of 2HG measurement in non-tumor tissue. Therefore, we tested how well TE-optimized 2HG levels distinguish between IDH-mutated glioma and non-tumor tissue, in this case, normal-appearing brain. We further examined the impact of different spectral modeling strategies (baseline stiffness, macromolecule inclusion, and basis set composition).

**Materials and Methods:** 48 patients with diagnosed/suspected IDH-mutated glioma were enrolled. 3T MRS data were acquired from tumor and contralateral non-tumor tissue with PRESS localization (TE = 97 ms, optimized for 2HG detection) and analyzed with ‘LCModel’ software. Receiver operating characteristic analysis evaluated 2HG estimates’ ability to distinguish IDH-mutated glioma from non-tumor brain tissue. Modeling interactions between 2HG and other metabolites were evaluated to identify reasons for potential false-positive 2HG detection.

**Results:** TE-optimized PRESS distinguished IDH-mutated glioma from non-tumor tissue with lower sensitivity (range 0.76-0.62) and specificity (0.85-0.78) than literature suggests for IDH-mutated vs. IDH-wildtype glioma. Strong negative correlations between gamma-aminobutyric acid (GABA) and 2HG persisted across all modeling strategies and may lead to false-positive 2HG detection in non-tumor tissue. We further present a cautionary example from a patient on a ketogenic diet, showing that the ketone body acetone can interfere with 2HG detection.

**Conclusions:** Spectral overlap with GABA and acetone can lead to false-positive 2HG detection in non-tumor tissue. Clinicians need to be mindful of these pitfalls when interpreting 2HG estimates.

## Introduction

Metabolic reprogramming is a hallmark of glioma. Key somatic mutations occur in the genes encoding isocitrate dehydrogenase 1 and 2 (IDH1/2).^1^ Gliomas with IDH mutations (IDHmut) display distinct biological behaviors compared to IDH wildtype (IDHwt). The presence of IDH mutations is associated with a more favorable outcome in glioma, including longer progression-free and overall survival, and increased sensitivity to treatment.^2^ IDH mutation status has therefore become one of the most critical factors in classifying gliomas.^3^

One of the key molecular alterations in IDHmut glioma is the production of the oncometabolite D-2-hydroxyglutarate (2HG) instead of α-ketoglutarate. In healthy brain tissue, 2HG is an error product of normal metabolism and therefore only present at negligible levels.^4,5^ However, in IDHmut gliomas, 2HG can accumulate at millimolar levels. At these concentrations, 2HG profoundly influences tumor biology by modulating cell death, epigenome, and metabolic processes.^6^ Consequently, targeting this neo-enzymatic activity with IDH inhibitors has emerged as a novel therapeutic class, with IDH1/2 inhibitor vorasidenib gaining FDA approval in 2024 for the treatment of gliomas.^7,8^

At pathological levels, 2HG can be measured non-invasively with *in-vivo* ^1^H magnetic resonance spectroscopy (MRS).^9^ MRS-derived estimates of 2HG may serve as useful biomarker for diagnosis and treatment monitoring in IDHmut glioma.^9–13^ MRS may be especially valuable in cases where a biopsy is considered too risky or not feasible. The key challenge in unequivocally measuring 2HG is the poor spectral dispersion at clinical field strength (i.e., 3T), which causes its three J-coupled multiplet signals (1.9, 2.25, 4.02 ppm) to overlap with signals from other metabolites, including glutamate (Glu), glutamine (Gln), and g-aminobutyric acid (GABA). The most commonly used approach is an optimized timing of the point-resolved spectroscopy (PRESS) localization sequence (TE/TE1/TE2 = 97/32/65 ms), originally proposed by Choi et al.^9^ This setup is designed to achieve a ‘pseudo-singlet’ appearance of the key resonances of Glu and 2HG, which reduces overlap in the 2.25 ppm region. Advanced MRS techniques (e.g., edited MRS^14^ and 2D correlation MRS^11^) have also been proposed for 2HG detection, but their lack of commercial availability, higher technical demands, vulnerability to measurement instabilities, and longer acquisition times have positioned conventional MRS sequences with optimized TE spacing as more viable for clinical translation^15^, although they are currently applied only in clinical research.^13^

A recent meta-analysis concluded that TE-optimized PRESS has high diagnostic specificity (97%) and sensitivity (92%) for IDHmut glioma. However, most studies that have assessed diagnostic performance to date have done so featuring IDHwt glioma as a comparison.^13^ 2HG detection should ideally facilitate diagnosis of IDHmut glioma while excluding other (non- IDHwt) conditions, i.e., non-glial primary brain tumors, inflammatory diseases, infections, infarction, and brain metastases.^15^ Notably, brain tumors exhibit very different spectral than normal brain, and other brain diseases. Specifically, they typically contain lower levels of the key metabolites that overlap with 2HG, like Glu and GABA^16^, which is likely to make unequivocal 2HG identification easier. To fully characterize the diagnostic accuracy of TE-optimized PRESS, one must also consider its performance in non-tumor tissue.

Furthermore, although all studies included in the abovementioned meta-analysis employed the widely used LCModel software for spectral analysis, they vary considerably in the details of the modeling strategies. It is established that important modeling decisions (such as baseline modeling and basis-set composition) influence metabolite estimation^17,18^, but this phenomenon has not been thoroughly investigated in the context of 2HG detection.

In this study, potential causes of false-positive 2HG detections in non-tumor tissue using TE-optimized PRESS are investigated. The performance of TE-optimized 2HG detection in differentiating IDHmut glioma from viable normal-appearing brain tissue from the contralateral hemisphere was systematically investigated and showed that modeling interactions between GABA and 2HG signals decrease diagnostic performance against normal-appearing tissue. Further, the effect of different spectral modeling strategies on performance was investigated, focusing on the impact of baseline modeling stiffness, macromolecule inclusion, and basis set composition. Finally, a cautionary example is presented which demonstrates that the presence of the ketone body acetone can interfere with 2HG detection.

## Methods

### Study Design

The study protocol was approved by the local institutional review board (IRB protocol NA_00036313), and written informed consent was obtained from all subjects. Subjects with IDHmut glioma diagnosis or suspected low-grade glioma at the time of the MRS scan, sufficient tumor volume (> 8 ml) estimated from 3D T2-weighted FLAIR images, and age >18 years were recruited. Datasets were only included in the subsequent analysis if IDHmut diagnosis was confirmed following biopsy/surgery (see Consort Diagram in Supplemental Data). IDH mutation status was determined by immunostaining (Anti-IDH1 R132H antibody) or next-generation sequencing (Johns Hopkins Hospital NGS Solid tumor panel).^19^

### Magnetic Resonance Acquisition

Patients were scanned using an MRS protocol on a 3T Philips Ingenia Elition scanner (Philips Healthcare, the Netherlands) with a 32-channel head coil. The protocol included 3D FLAIR, 3D T1-weighted MPRAGE, B_0_ map for shimming^20^ and ^1^H single-voxel spectroscopy.

MR spectra were acquired using the vendor-provided product PRESS sequence at two different TEs, 30 ms (128 transients) and 97 ms (256 transients), with TR = 2 s and the same B_0_ shimming parameters. Echo spacing in PRESS at 97 ms was optimized to detect the 2HG signal at ∼2.25 ppm (TE1/TE2 = 32/65 ms).^9^ (See Supplemental Data for MRSinMRS Reporting Checklist^21^).

The spectroscopic voxel was placed in the T_2_-hyperintense tumor area defined on FLAIR images. Voxel size was adjusted according to tumor size to reduce partial volume effects (8-27 ml). In the presence of a large tumor, data were acquired from multiple locations within the lesion in several participants. Control spectra were obtained from anatomically similar contralateral, normal-appearing brain regions. Water-unsuppressed spectra were acquired from the same volumes of interest at the same TR and TE (1 transient) for water-referenced metabolite quantification and eddy-current correction.

### Spectral Analysis

MRS data were analyzed using the open-source toolbox Osprey (v. 2.6.3; https://github.com/schorschinho/osprey/)^22^ , using the integrated LCModel binary for fitting.^23^ Data had already been coil-combined, averaged and eddy-current-corrected on the scanner. Basis sets were simulated with MRSCloud^24^, assuming ideal excitation, with sequence timings and refocusing RF pulse waveforms used in the actual sequence and effects of spatial localization considered (30 × 30 × 30 mm^3^ volume, 41 x 41 points). The basis set included 2HG, alanine (Ala), ascorbate (Asc), aspartate (Asp), citrate (Cit), creatine (Cr), negative creatine methylene (-CrCH2), cystathionine (Cystat), ethanolamine (EA), GABA, glycerophosphocholine (GPC), glutathione (GSH), glucose (Glc), Gln, Glu, glycine (Gly), myo-inositol (mI), lactate (Lac), N-acetylaspartate (NAA), N-acetylaspartylglutamate (NAAG), phosphocholine (PCh), phosphocreatine (PCr), phosphoethanolamine (PE), scyllo-inositol (sI), serine (Ser), taurine (Tau).^25^ If participants were following a ketogenic diet (see Results section “Strong interference of acetone with 2HG warrants caution in patients on the ketogenic diet”), acetoacetate (AcAc), acetone (Ace), and β-hydroxybutyrate (β-OHB) were included. The spectral fitting range was 0.5-4.1 ppm, including all 2HG resonances. Full-width at half-maximum (FWHM) estimates were used for spectral quality assessment; spectra with FWHM >12 Hz were excluded.^26^

### Modeling Strategies

Spectral modeling was repeated with seven baseline stiffness settings (varying the spline knot spacing parameter ‘DKNTMN’ from 0.15 to 5). Modeling was further performed with and without LCModel-simulated macromolecules and lipids (MM09, MM12, MM14, MM17, MM20, Lip09, Lip13a, Lip13b, Lip20).

To evaluate the goodness of fit in the target spectral region (2.0-2.4 ppm), the fit quality number (FQN, recommended by expert consensus) was calculated by dividing the fit residual variance by the pure measurement noise variance .^27^ We hypothesized that false-positive 2HG modeling would lead to subtle modeling imperfections, which would result in higher fit residual and, consequently, be identifiable through elevated FQN.

### Metabolite Quantification

Metabolite measurements were quantified relative to the internal tissue water signal. Water T_2_ relaxation correction was performed using T_2_ estimates derived by fitting a linear model to the logarithm of the water signal intensities against the two available TEs, 30 ms and 97 ms^28^, (see Supplemental Data for T_2_ estimates). SPM12^29^ was used to perform tissue segmentation to yield tissue volume fractions of GM, WM, and CSF in normal-appearing brain, and water concentration was calculated using tissue-specific water concentrations from literature.^30^ For the tumor tissue, we assumed an equal composition of gray and white matter and used a water concentration value of 42.3 M, as proposed by Choi et al.^9^ To evaluate the performance of dual-TE T_2_ estimation, metabolite estimates in normal-appearing brain tissue were re-calculated using tissue-specific water T_2_ given in the literature^30,31^ and presented in Supplemental Data.

### Statistical Analysis

2HG estimates in IDHmut tumor and contralateral normal-appearing brain were compared using non-parametric Wilcoxon rank sum testing following the Shapiro-Wilk test confirming non-normality. Two outcome measures were used to investigate the relationship between estimates of 2HG and other metabolites. First, LCModel coefficients of model covariance (CMC) were extracted, a measure of interaction between two model parameters *within a single dataset*, derived from the Fisher information matrix. Second, correlations between 2HG and other metabolite estimates across contralateral MRS datasets were assessed by Spearman analysis, following Shapiro-Wilk testing confirming non-normality.

Receiver operating characteristic analysis was performed to evaluate 2HG estimates’ ability to distinguish IDHmut glioma and non-tumor brain tissue. Detection of 2HG in non-tumor spectra obtained from normal-appearing brain was considered a false-positive. The optimal decision threshold for 2HG was calculated using the Youden Index , which provides the threshold that maximizes the difference between the true positive rate (sensitivity) and the false positive rate (1 - specificity).

To investigate the association between fitting goodness and false-positive 2HG detection in control spectra, FQN and 2HG estimates were correlated using the Spearman correlation test. Furthermore, an additional ROC analysis was performed to assess the performance of FQN in distinguishing false-positive 2HG detection. All analyses were performed using R v4.4.1 in RStudio v2024.04.2. Results were considered significant at p < 0.05.

To validate the dual-TE quantification approach, Spearman correlation analysis was performed between metabolite estimates in normal-appearing brain tissue calculated using dual-TE water T_2_ estimates and tissue-specific literature-based water T_2_ relaxation times. Water T_2_ estimates of IDHmut tumor and normal-appearing contralateral tissue in MRS voxels were compared using the Wilcoxon rank-sum test, following confirmation of non-normality.

## Results

### Patient Characteristics

Forty-eight glioma patients were prospectively enrolled and underwent the MRI/MRS examination. Six patients with undefined histological status and two patients diagnosed with IDHwt glioblastoma were subsequently excluded (see Consort Diagram in Supplemental Data). Twenty-four of the remaining forty patients were diagnosed with IDHmut astrocytoma and sixteen with IDHmut oligodendroglioma. Thirty-seven patients underwent partial resection and had measurable residual disease before the MRS examination. Thirteen patients had a history of radiotherapy and chemotherapy (no IDH inhibitor). Patient characteristics are listed in **Table 1**.

**Table 1.** Patient characteristics.

| **Characteristics** | **All IDHmut patients**<br>**(n = 40)** |
| --- | --- |
| ***General*** |  |
| Age, median (range) | 40 (18-79) |
| Female | 12 |
| ***Histology*** |  |
| Oligodendroglioma, IDH-mutant, 1p/19q-codeleted, grade 2 | 12 |
| Oligodendroglioma, IDH-mutant, 1p/19q-codeleted, grade 3 | 4 |
| Astrocytoma IDH-mutant, grade 2 | 17 |
| Astrocytoma IDH-mutant, grade 3 | 7 |
| ***IDH mutation status*** |  |
| IDH1 R132H-mutated | 37 |
| IDH1 R132C-mutated | 2 |
| IDH2 R172K-mutated | 1 |
| ***Prior treatment*** |  |
| Resection | 37 |
| Radiotherapy | 13 |
| Chemotherapy | 13 |

In twenty-eight cases, control spectra were obtained from the contralateral normal-appearing brain as described above (**Figure 1A-B**). In six IDHmut cases, data were acquired from multiple locations (2-4) within the lesion. Based on data exclusion criteria (FWHM >12 Hz), one tumor spectrum and one control spectrum were excluded, leaving 47 IDHmut tumor spectra and 27 contralateral spectra for further analysis.

**Figure 1.**
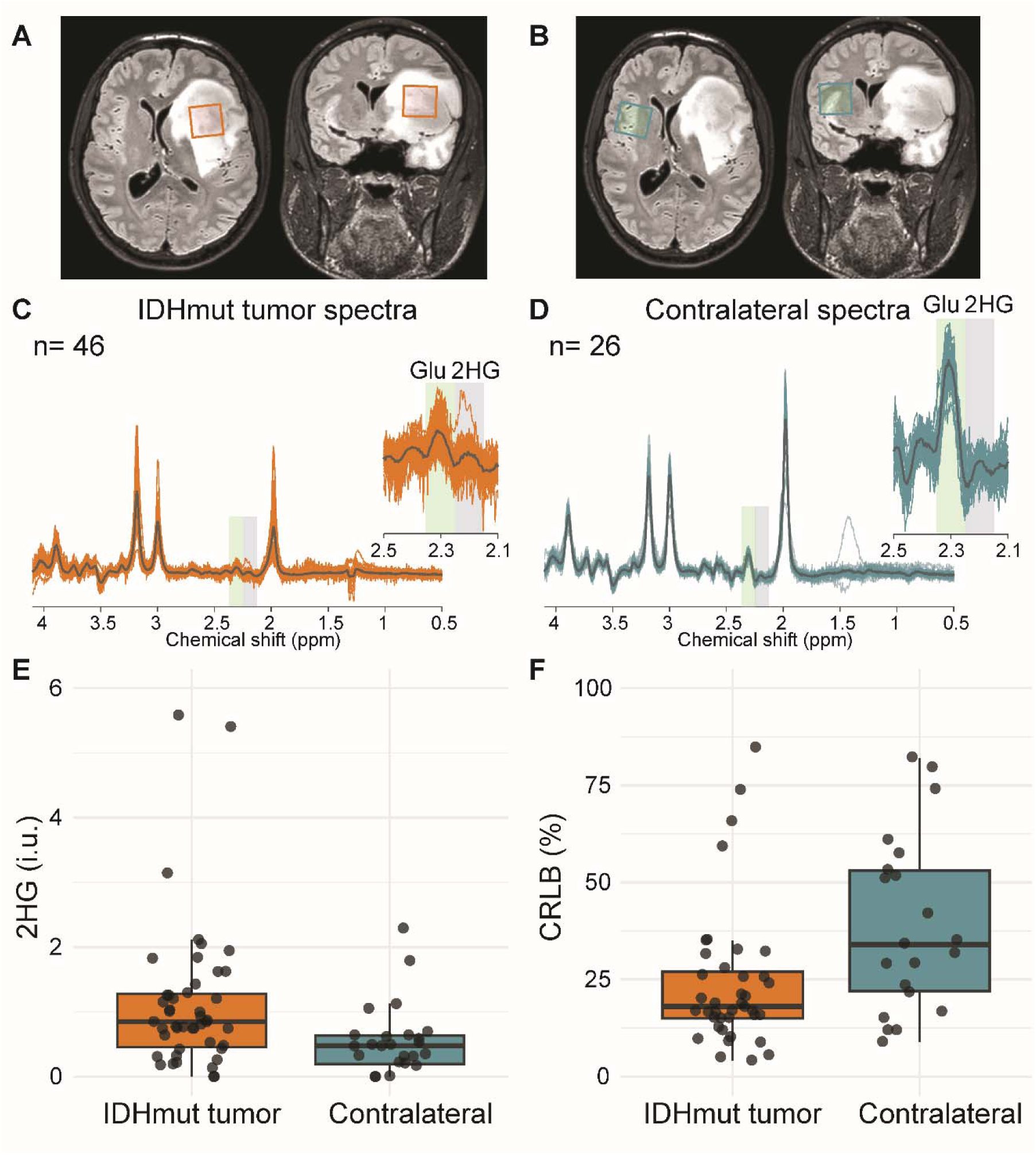
Example voxel locations in tumor (A) and contralateral hemisphere (B) in one subject overlaid on FLAIR MRI. In-vivo MR spectra acquired from (C) IDHmut glioma (n = 46) and (D) contralateral brain (n = 26) using TE-optimized PRESS at 3T for 2HG detection. One additional IDHmut tumor and contralateral spectra are shown in Figure 5 and are not included here. Mean spectra are displayed in black. The spectral regions with major peaks of Glu and 2HG are magnified and highlighted with green and gray boxes. Boxplots of (**E)** 2HG estimate and (**F**) spectral fitting uncertainty values (CRLB) obtained from LCModel fitting of IDHmut tumor and contralateral spectra. Thirteen data points (seven IDHmut, six contralateral) with CRLB >100% are not shown.

### 2HG estimates in IDHmut tumor and normal-appearing brain tissue

Characteristic patterns in the 2.1-2.5 ppm region highlight lower Glu and higher 2HG signals in IDHmut spectra than contralateral spectra (**Figure 1C-D**). Despite this clear visual distinction, there was no obvious 2HG cut-off that would perfectly separate the tumor and contralateral groups (**Figure 1E**), although median 2HG estimates were significantly higher in tumor (median 2HG_tumor_ = 0.85 i.u. and median 2HG_control_ = 0.47 i.u., p = 0.001). As shown in **Figure 1F**, 2HG was detected in seven control spectra with high spectral fitting certainty (i.e., Cramér–Rao lower bound (CRLB) < 25%).

The correlation between 2HG estimates in the contralateral voxel calculated using dual-TE T_2_ estimates and those calculated using tissue-specific water T_2_ relaxation times (segmentation-based) was nearly perfect (ρ = 0.99) (see Supplemental Data). Water T_2_ estimates in IDHmut tumor voxels (114.0 ± 32.8 ms) was significantly higher than in normal-appearing contralateral voxels (67.9 ± 3.61 ms) (p < 10^-16^), consistent with literature.^32^ This confirmed that it is justified to use 2HG estimates in both tumor and contralateral calculated using dual-TE T_2_ estimates.

### Interactions between 2HG and other metabolites’ spectral models

Strong spectral overlap of 2HG and GABA between 2.1-2.5 ppm (**Figure 2A**) resulted in large negative model covariance between the two metabolites in both tumor and non-tumor spectra (median CMC_tumor_ = -0.44 and median CMC_control_ = -0.49), suggesting it remains difficult for the modeling process to separate them reliably (**Figure 2B**). 2HG estimates interacted similarly, but more weakly, with tCr, Glu, tNAA, Glx, NAAG, MM20, PCr and tCho. Interactions between MM20 (macromolecules at 1.95, 2.0, 2.25 and 3.0 ppm) and 2HG were more prominent in IDHmut tumor spectra than contralateral spectra (median CMC: -0.12 and -0.03, respectively).

**Figure 2.**
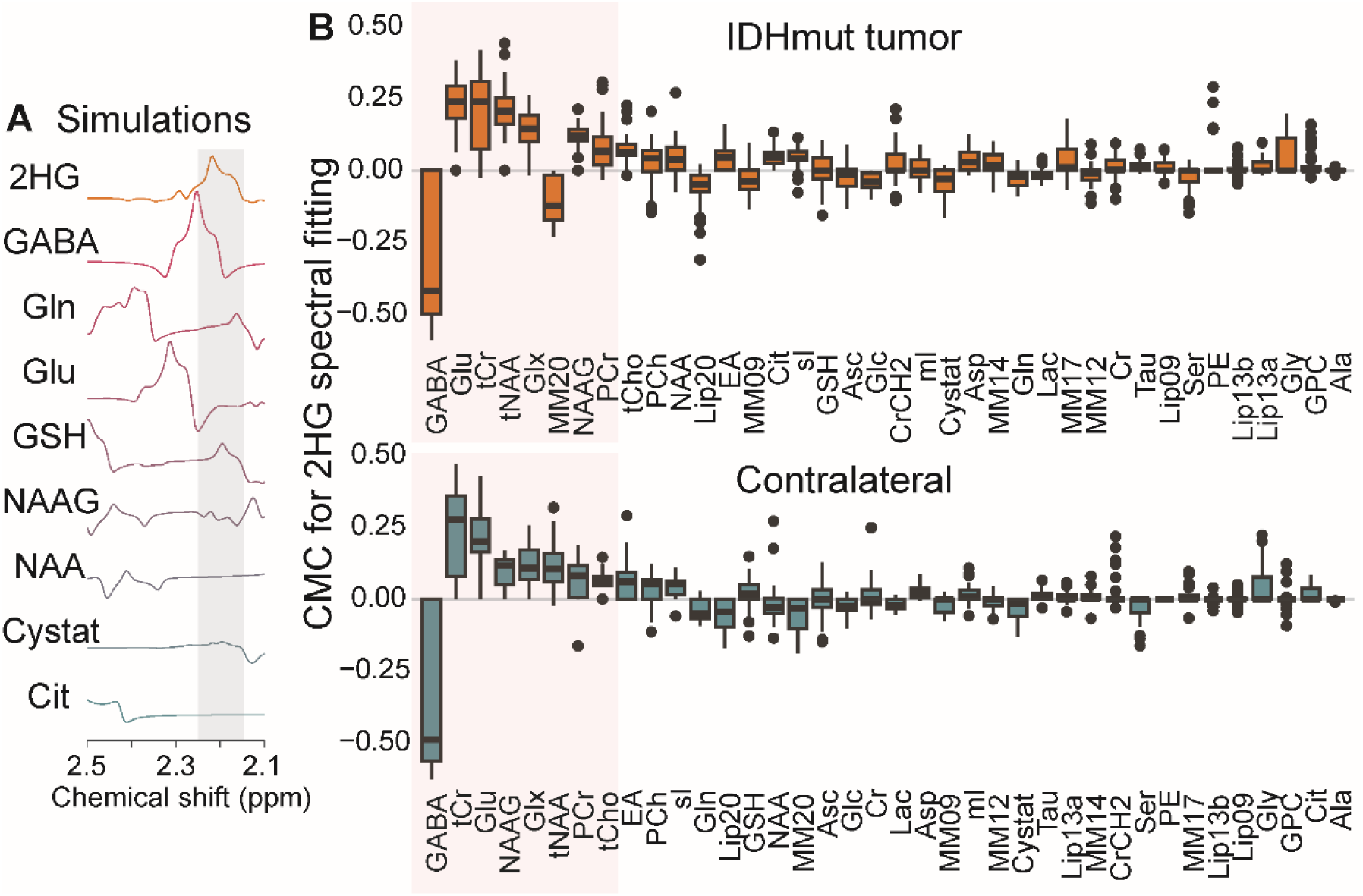
**(A)** Spectral simulations of metabolites that have resonances in the same spectral region as 2HG (2.1-2.5 ppm). The spectral region with major peaks of 2HG is highlighted with a gray box. (**B)** Coefficients of modeling covariance (CMC) between 2HG and other small metabolites and macromolecules, calculated by LCModel for IDHmut tumor and contralateral control spectra. Boxplots display the inter-quartile range (IQR) with whiskers extending 1.5’IQR, along with median lines and outlier points. The boxplots are ordered by absolute median CMC values. The largest (negative) CMC values were observed with GABA for both spectrum types, which also exhibits the greatest spectral overlap with 2HG.

The strong negative association between 2HG and GABA was again present in the correlation of their amplitude estimates across all contralateral datasets (p < 10^-5^, ρ = -0.8) (**Figure 3**). There was no significant correlation between 2HG and other metabolite estimates in the contralateral data.

**Figure 3.**
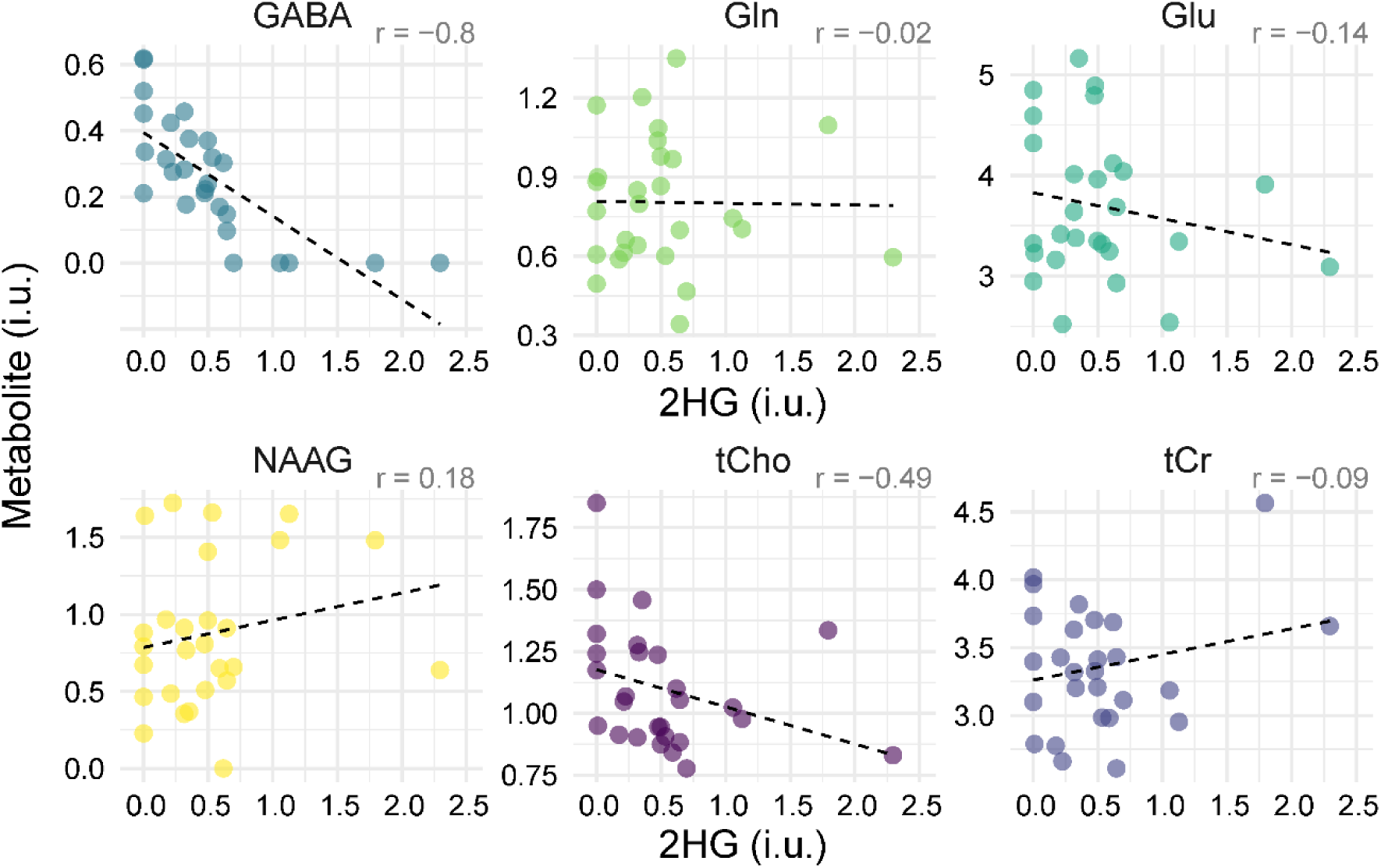
Correlation between estimates of 2HG concentration and other metabolites (which have high CMC, see Figure 2) in contralateral spectra. Presumed false positive detection of 2HG in normal-appearing brain tissue is inversely correlated with GABA levels (p < 0.00001).

### Influence of baseline and macromolecule modeling on 2HG estimates

The negative 2HG and GABA association persisted strongly across different modeling strategies, independent of the choice of baseline stiffness and inclusion of macromolecular profiles in the basis set (p < 10^-4^ to 10^-7^, ρ = -0.7 to -0.83) (**Figure 4**), although mean 2HG levels decreased considerably with increasing baseline flexibility across all datasets (mean 2HG_tumor_ = 1.11 i.u. to 1.02 i.u.; mean 2HG_control_ 0.51 i.u. to 0.33 i.u.), suggesting that flexible baseline models absorb signal from 2HG. Although MRS estimates of GABA are assumed to be higher in GM than in WM^33^, no association was observed between contralateral brain tissue composition (i.e., GM content) and 2HG and GABA estimates (see Supplemental Data).

**Figure 4.**
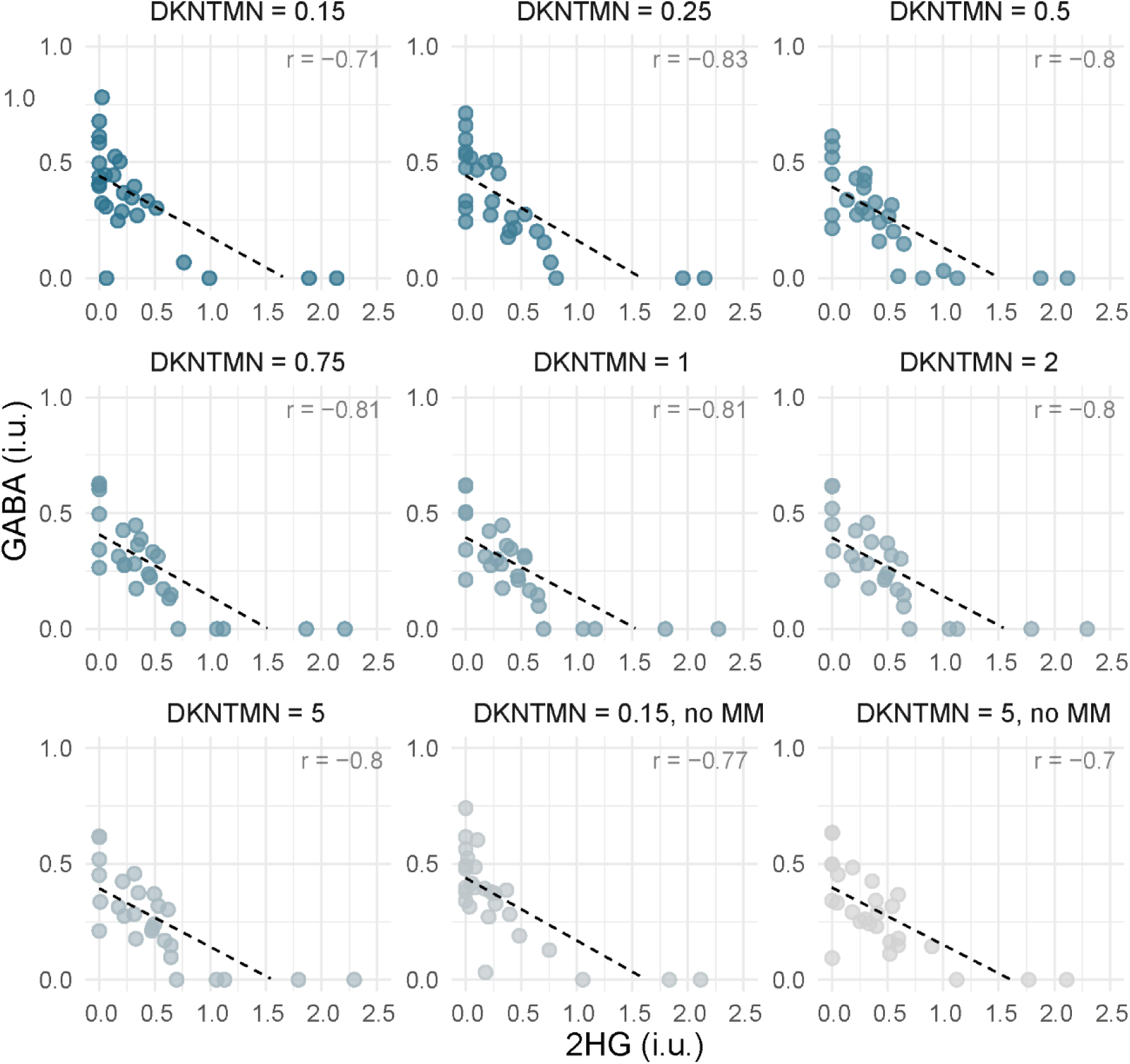
Correlation between GABA and 2HG estimates across various baseline stiffness (defined with DKNTMN control parameter) and with/without inclusion of macromolecule (MM) signals in LCModel spectral fitting. DKNTMN values correspond to the minimum spacing between spline knots. The inverse correlation between GABA and 2HG detection remained highly significant across all analyses (p < 0.001). The blue-to-gray gradient encodes DKNTMN.

The area under the ROC curve (AUC), a measure of diagnostic accuracy, was highest (0.78) for 2HG estimates obtained with flexible baseline modeling (DKNTMN = 0.15) (**Figure 5**). The optimum cutoff 2HG level for this case was 0.44 i.u., with a sensitivity of 0.79 and specificity of 0.81. The range of AUCs across all modeling strategies was 0.78–0.69, with an optimum threshold for 2HG estimates between 0.42–0.72 i.u., sensitivity of 0.76–0.62, and specificity of 0.85–0.78.

**Figure 5.**
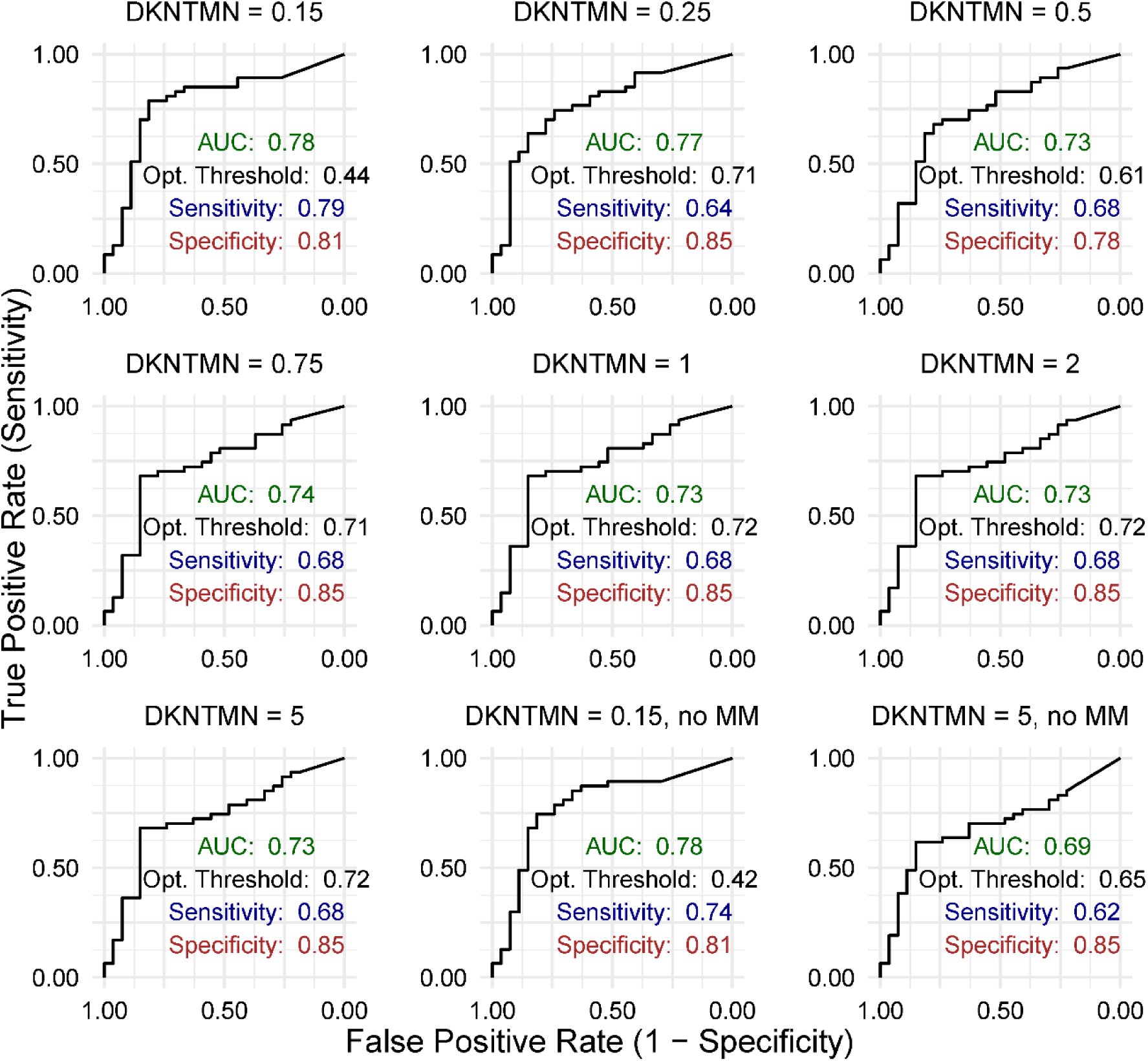
Receiver operating characteristic curves showing the performance of 2HG estimates in distinguishing IDH-mutant glioma from non-tumor tissue for various spectral modeling approaches (i.e., various baseline stiffness defined using DKNTMN control parameter and with/without the inclusion of macromolecule, MM, signals in spectral fitting). Each plot includes the area under the curve (AUC), along with sensitivity, and specificity values for the optimum threshold for 2HG estimates.

When the stiffest baseline modeling approach was applied, excluding macromolecule signal models (DKNTMN = 5, no MM), FQN for the 2–2.4 ppm region (targeting the 2HG peak at ∼2.25 ppm) showed good performance (AUC = 0.71) in identifying false-positive 2HG detections in control spectra. The optimal cutoff for FQN was <1.4, yielding a sensitivity of 1 and a specificity of 0.57 (see Supplemental Data).

### False-positive 2HG detection in a participant on a ketogenic diet

Finally, a strong overlap between 2HG and the acetone singlet at 2.22 ppm can cause false-positive 2HG detection (**Figure 6**). Unusually high 2HG levels were quantified in both tumor and contralateral spectra of one IDHmut glioma patient who was on a ketogenic diet. Therefore, the dataset was re-analyzed with the ketone bodies, acetone, acetoacetate, and b-hydroxybutyrate included in the basis set. After inclusion, acetone and b-hydroxybutyrate were fitted in both spectra, while 2HG was estimated substantially lower in tumor, and not at all in contralateral. The high CMC (-0.69) between 2HG and acetone confirmed strong modeling interaction between these two compounds.

**Figure 6.**
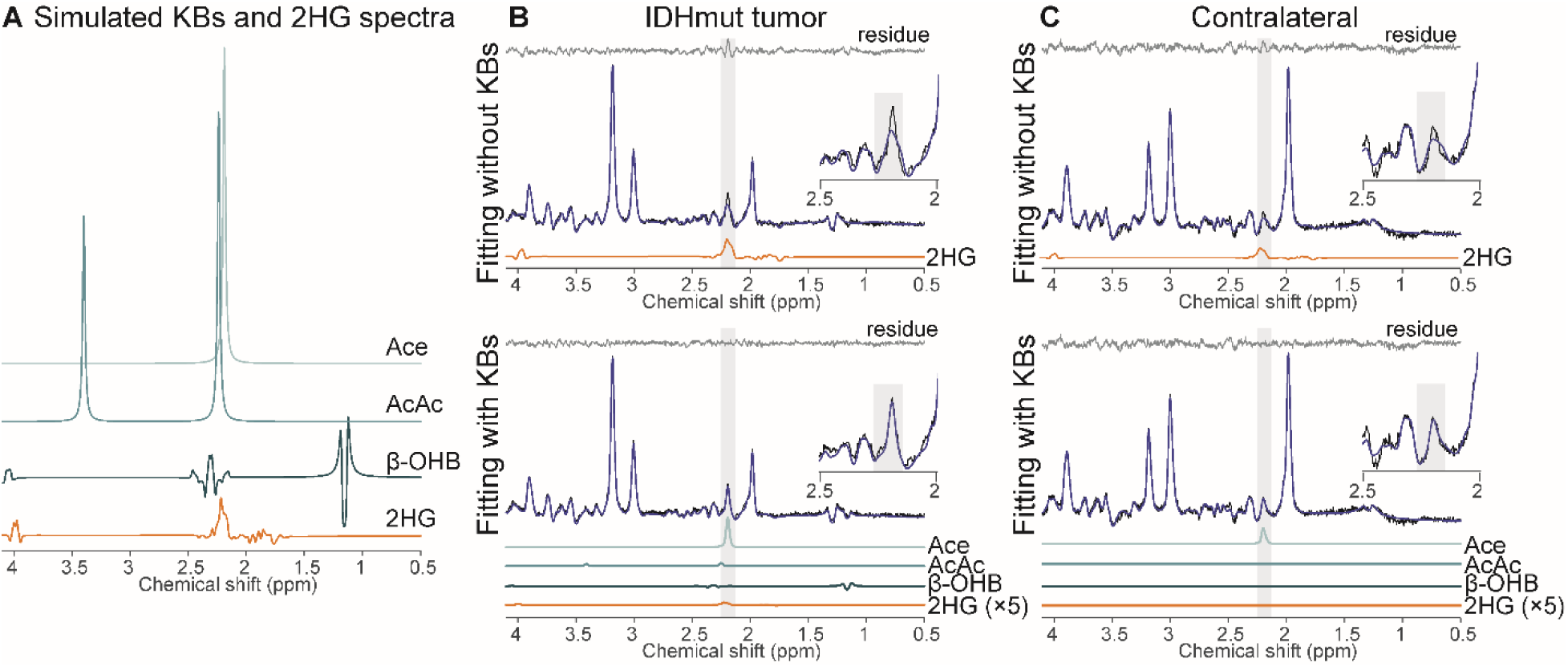
Example of false-positive high-level 2HG detection in both the tumor and normal-appearing contralateral brain tissue of an IDHmut glioma patient following a ketogenic diet. (**A)** Simulated spectra of acetone (Ace), acetoacetate (AcAc), beta-hydroxybutyrate (b-OHB), and 2HG. Fitting of (**B**) IDHmut tumor and (**C**) contralateral spectra with/without ketone bodies, strongly influencing 2HG estimates. Spectral data are presented in black and fitting models in dark blue. Individual fitting models are shown for 2HG and ketone bodies.

## Discussion

### TE-optimized MRS can give false-positive 2HG findings in non-tumor tissue

The current study presents evidence that positive 2HG findings on TE-optimized MRS do not mean an unequivocal IDHmut glioma diagnosis, since a 2HG signal was quantified in normal brain with low uncertainty values in 7 of 27 control spectra. We speculate this might also be found in other lesion types such as infection, inflammation, or others, depending on their GABA levels. Understanding limitations and failure modes of 2HG MRS is especially critical for cases where biopsy is not feasible and MRS would be the only means of diagnosis. Our data show that 2HG estimates from TE-optimized PRESS can be artifactually higher than expected in spectra from viable non-tumor tissue with normal MRI appearance. The performance of TE-optimized PRESS in distinguishing IDHmut glioma from contralateral normal-appearing tissue was therefore noticeably lower compared to its ability to differentiate IDHmut from IDHwt glioma.^13^ This discrepancy can be attributed to low GABA concentrations in both IDHwt glioma and IDHmut^34^, potentially causing the 2HG-GABA association to be overlooked.^35^

2HG detection performance has not been systematically investigated in pathologies that may mimic glioma. This literature gap may give clinicians a false sense of diagnostic certainty of 2HG MRS, particularly if those lesions have normal or elevated GABA levels. Furthermore, the risk of false-positive 2HG detection likely increases if the measurement volume contains appreciable amounts of normal brain tissue, e.g., with small and/or infiltrating lesions.

### GABA strongly interferes with 2HG estimation independent of modeling strategy

The primary cause appears to be spectral overlap not entirely eliminated by TE optimization. Coefficients of modeling covariance, a measure of interaction between model parameters during spectral fitting, revealed a strong anti-correlation between GABA and 2HG, supporting similar findings for sLASER localization at 3T and 7T.^36^ This increases the likelihood of the GABA signal being erroneously modeled as 2HG. The strong negative correlation between 2HG and GABA estimates in contralateral tissue gives further validation. While this study used data from normal-appearing contralateral brain, our findings suggest that 2HG MRS in lesions with normal (or high) GABA levels may lead to false-positive 2HG detection, which may, in turn, lead to false diagnoses if MRS were considered in isolation. Notably, 26% of contralateral spectra exhibited low 2HG fitting uncertainty (CRLB < 25%), although very little 2HG signal is expected in normal brain. The practice of using high relative CRLB values to identify low detection certainty and exclude data is commonly criticized for inducing bias^37^, but the current data conversely show that low CRLBs alone do not guarantee accurate 2HG detection. Nevertheless, CRLB cut-off criteria remain popular, and previous 2HG studies have proposed cut-offs for reliable 2HG detection as high as 50%^14,17^, which would include most contralateral datasets in our study.

It was also demonstrated that the strong GABA-2HG interaction persists across many different modeling strategies. Different modeling strategies can affect the estimation of metabolite levels, particularly for J-coupled signals like Glu, GABA, and 2HG. Indeed, changes in baseline modeling in our work influenced 2HG estimates and, consequently, the overall performance, including the optimal threshold for 2HG estimation, as well as diagnostic sensitivity and specificity. The GABA-2HG interaction remained unanimously strong, suggesting that the overlap problem cannot be solved at the modeling level.

Previous work demonstrated that the sensitivity of MRS for detecting 2HG depends on tumor volume, recommending 8 mL as lower limit for reliable detection^15^. Conversely, a small brain lesion could also lead to partial volume effects, since normal-appearing brain tissue included in the MRS voxel could increase GABA levels in the measurement volume and facilitate false-positive 2HG detection, for example in a non-IDHmut-glioma lesion, or lead to overestimation of 2HG concentration in IDHmut glioma. While this might plausibly lead to assuming that grey-matter-rich voxels are more likely to exhibit higher (falsely detected) 2HG levels due to their greater GABA content, we did not observe an association between false-positive 2HG detection and the GM content of the contralateral volume of interest. In searching for a reliable indicator of false-positive 2HG detection, a quantitative estimate of the modeling quality, i.e., FQN specific to the 2HG region (2-2.4 ppm), was investigated. Indeed, the FQN was reasonably good in identifying false-positive 2HG detection (AUC = 0.71), but only for the stiffest baseline approach without the inclusion of macromolecule models..

### Weaker interference of other metabolites with 2HG

Other signals interacting with 2HG include Glu, Gln, NAAG, and MM20. The interactions did not vary much between tumor and control spectra except for MM20, which was more prominent in tumor spectra. Although there is no direct overlap between 2HG and tCr, their high coefficient of modeling covariance is likely again mediated through GABA, since GABA overlaps with 2HG at 1.9 and 2.25 ppm, and with tCr at 3 ppm, causing indirect interaction. These weaker modeling interactions did, however, not lead to negative correlations between 2HG and other metabolite estimates across all datasets in contralateral tissue.

### Strong interference of acetone with 2HG warrants caution in patients on ketogenic diet

Finally, this study showed that the acetone singlet (2.22 ppm) can also be mistakenly modeled as 2HG. This is highly important, as nutritional interventions (fasting, ketogenic diets) are often explored as complementary treatment approaches in low-grade glioma^38^. Ketone bodies (acetone, acetoacetate, β-hydroxybutyrate) are typically undetectable via conventional MRS in healthy brain under a standard diet, and therefore not commonly included in basis sets. However, in the context of nutritional interventions or specific dietary preferences, brain ketone concentrations can rise above the MRS detectability threshold, especially in brain tumors^39,40^. Therefore, the basis set composition needs to be modified based on medical history. High covariance between 2HG and acetone models underscores the difficulty in reliably separating them. Including acetone in the basis set *by default* may lead to *underfitting* of the 2HG signal, as it may reasonably assume some of the real-existing 2HG signal. Data-driven determination of basis set composition has been offered as an alternative to conventional basis sets^41^ In general, for study cohorts or clinical cases involving non-standard diets, the TE-optimized PRESS sequence may not be the ideal method for unequivocal 2HG quantification.

### Other 2HG MRS detection techniques

*J*-difference editing of 2HG may provide higher sensitivity and specificity than conventional protocols like the TE-optimized technique used in this work^14^. However, spectral editing is more technically demanding, vulnerable to experimental instabilities (motion or field drift), requires more nuanced post-processing, and is less widely available on clinical MRI scanners. Conventional approaches are likely to remain relevant, making it important to understand their failure modes. sLASER localization (TE = 110 ms^42^) and triple-refocusing^35^ attempt to further reduce spectral overlap between 2HG and metabolites, including GABA, Glu and Gln, but like TE optimization, they cannot remove it entirely, and likely experience the same problematic interaction with GABA. 2D correlation MRS techniques have also been employed to detect 2HG signals.^11^ However, like spectral editing, adoption in clinical practice is limited, owing to long acquisition times, demand for complex postprocessing, and limited availability on clinical scanners.

### Limitations

Our study is limited by a small sample size (n = 40). However, it is the largest high-quality TE-optimized PRESS dataset to date to compare spectra from IDHmut gliomas and contralateral normal-appearing brain. Due to the limited number of treatment-naïve patients, we combined pre- and post-treatment gliomas in our analysis,. 2HG levels may decrease after treatment (including chemoradiation), which could lower detection sensitivity in post-treatment patients.^43^ Additionally, water T_1_ and metabolite T_1_ and T_2_ relaxation time corrections were not performed, as tumor-specific relaxation times were unavailable. Nevertheless, we assumed that the major variability in relaxation correction originates from water T_2_ relaxation and calculated a specific T_2_ estimate using a dual-TE approach.

## Conclusions

TE-optimized MRS at 3T can exhibit false-positive 2HG detection in non-tumor tissue. Signal overlap with GABA and acetone (if patients are on a ketogenic diet/fasting) is the primary reason. Spectral modeling strategies such as baseline handling, macromolecule modeling, and basis set composition can furthermore influence the diagnostic accuracy of TE-optimized PRESS. In conclusion, 2HG estimates from TE-optimized PRESS should be interpreted with caution to determine IDH mutation status, considering possible overlap with GABA and acetone.

## Data Availability

MRS result tables and statistical analysis code that support the findings of this study are available from the corresponding author upon reasonable request.

## Supplementary material

Supplementary material is available.

## Abbreviations

2HG: 2-hydroxyglutarate
IDH: Isocitrate dehydrogenase
PRESS: Point-resolved spectroscopy
GABA: Gamma-aminobutyric acid
FQN: Fit quality number
CRLB: Cramér–Rao lower bound
CMC: Coefficients of modeling covariance

