## Supplementary Information for "Pitfalls in 2HG detection with TE-optimized MRS at 3T"

**Supplementary Information: Pitfalls in 2HG detection with TE-optimized MRS at 3T**

### Detailed Study Design

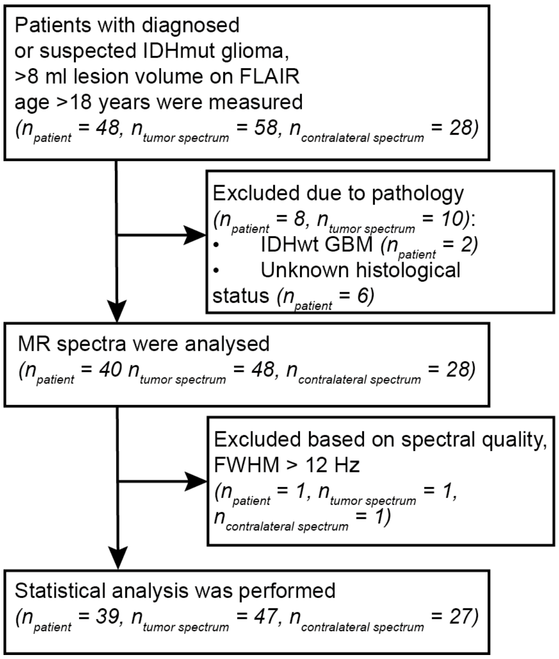


**Supp Figure 1.** Consort diagram showing patient recruitment with inclusion criteria and exclusion based on pathological findings and MR spectra quality.

### Metabolite quantification using dual-TE water reference

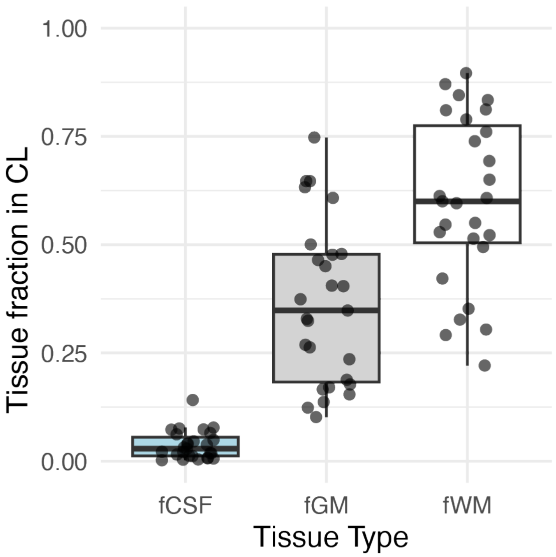


**Supp Figure 2.** Fraction of each tissue type (white matter, grey matter, cerebrospinal fluid) in contralateral voxels. The central line in each box denotes the median, the edges of the boxes represent the interquartile range (IQR), and the whiskers extend to 1.5 times the IQR.

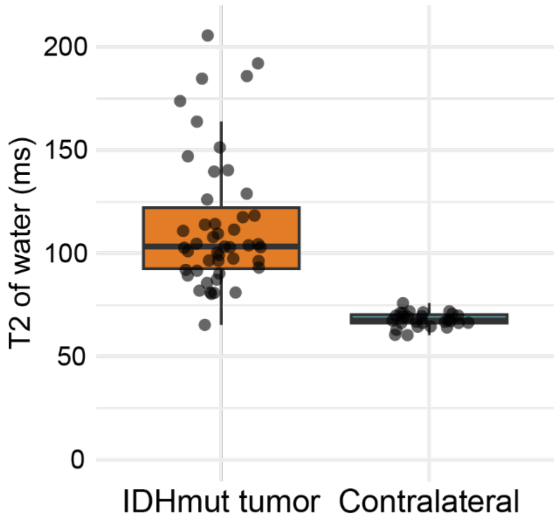


**Supp Figure 3.** Water T2 estimates of IDH-mutated tumor and contralateral normal-appearing brain tissue in MRS voxels. The central line in each box denotes the median, the edges of the boxes represent the interquartile range (IQR), and the whiskers extend to 1.5 times the IQR.

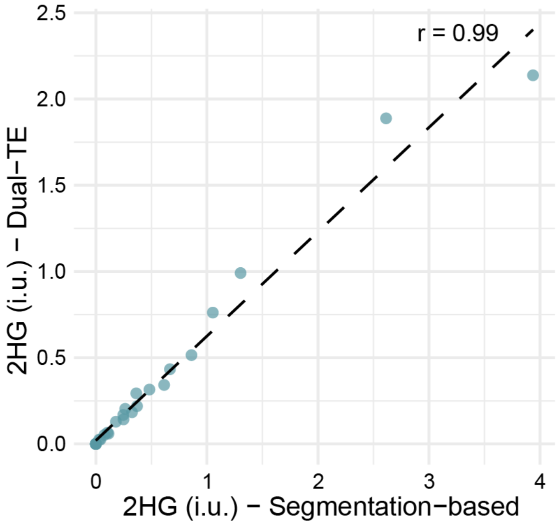


**Supp Figure 4.** Correlation plot between 2HG estimates in contralateral calculated using two different water T2 estimation approaches. The first approach (dual-TE) uses MRS water references acquired at two different TEs (30 ms and 97 ms) for water T2 estimation. In the second approach, literature-based tissue-specific along with tissue fractions were used for the water T2 calculation. A perfect correlation was observed between 2HG estimates calculated using these two techniques.

**Correlation between tissue fraction in normal-appearing brain and false-positive 2HG detection**

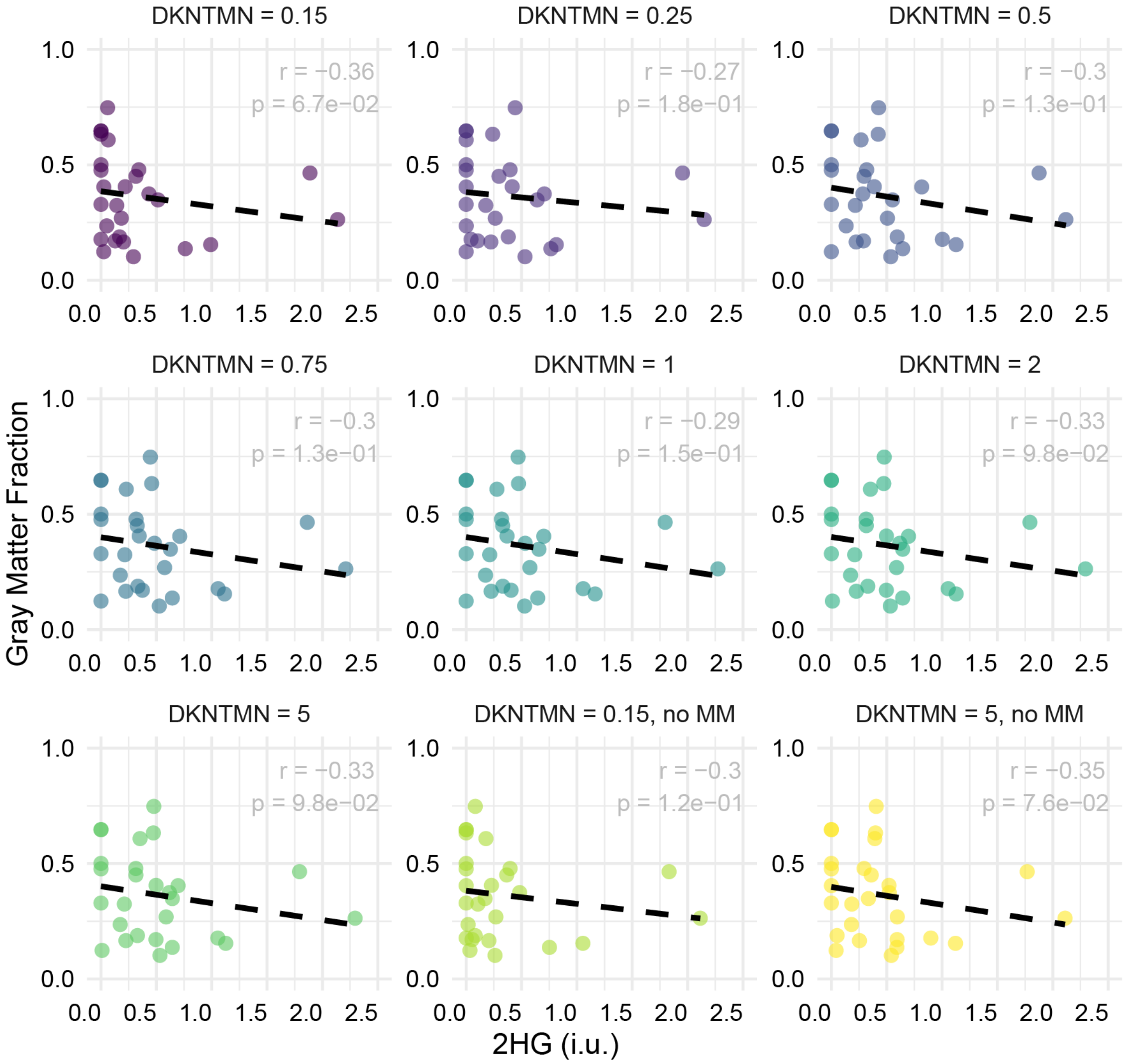


**Supp Figure 5.** Correlation analysis between gray matter fraction in contralateral MRS voxel and 2HG estimates across all modeling strategies. No significant correlation was observed.

**Association between fit quality number (FQN) and false-positive 2HG detection**

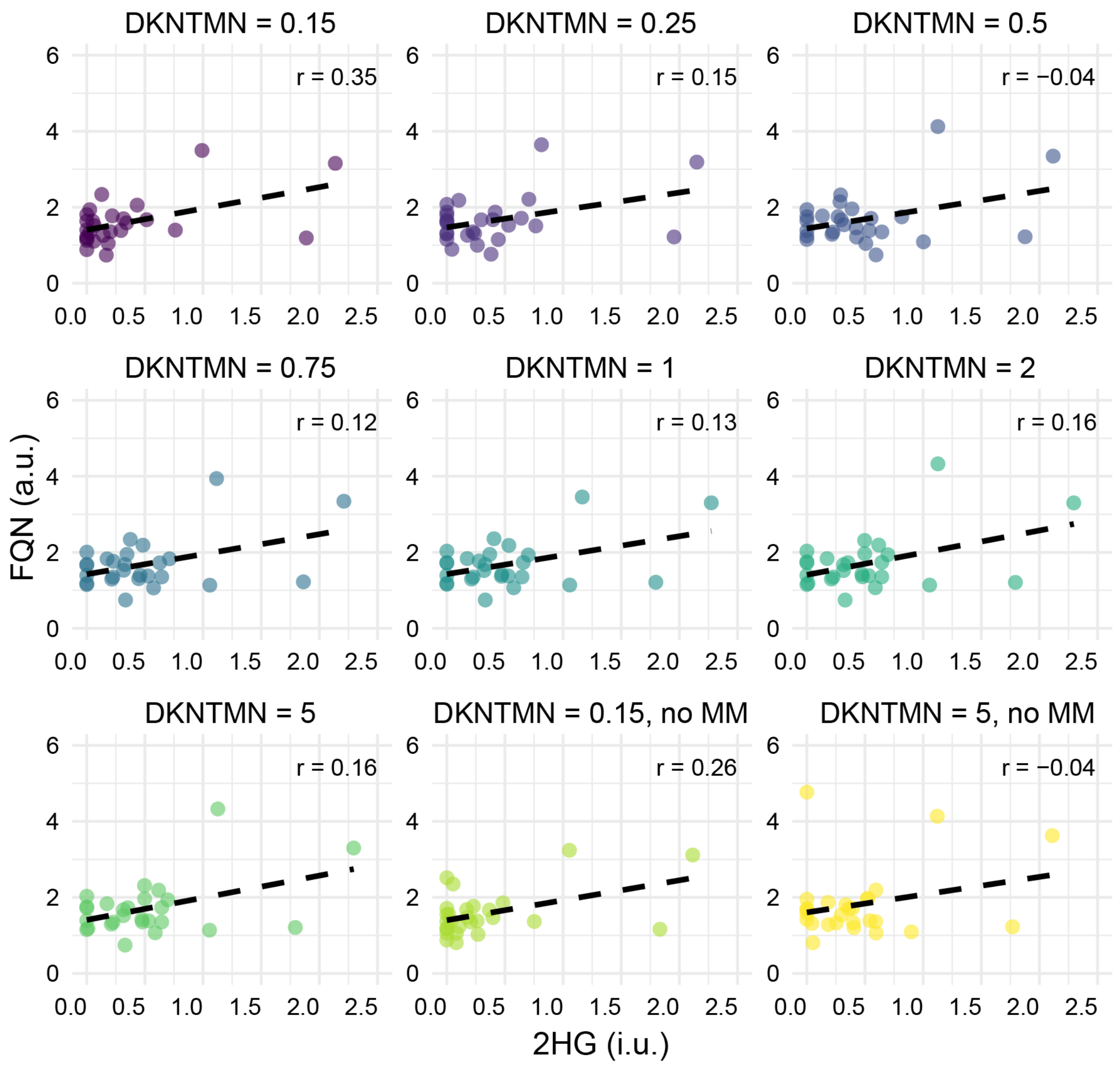


**Supp Figure 6.** Correlation analysis between fitting quality number (FQN) and 2HG estimates in contralateral across all modeling strategies. No significant correlation was observed.

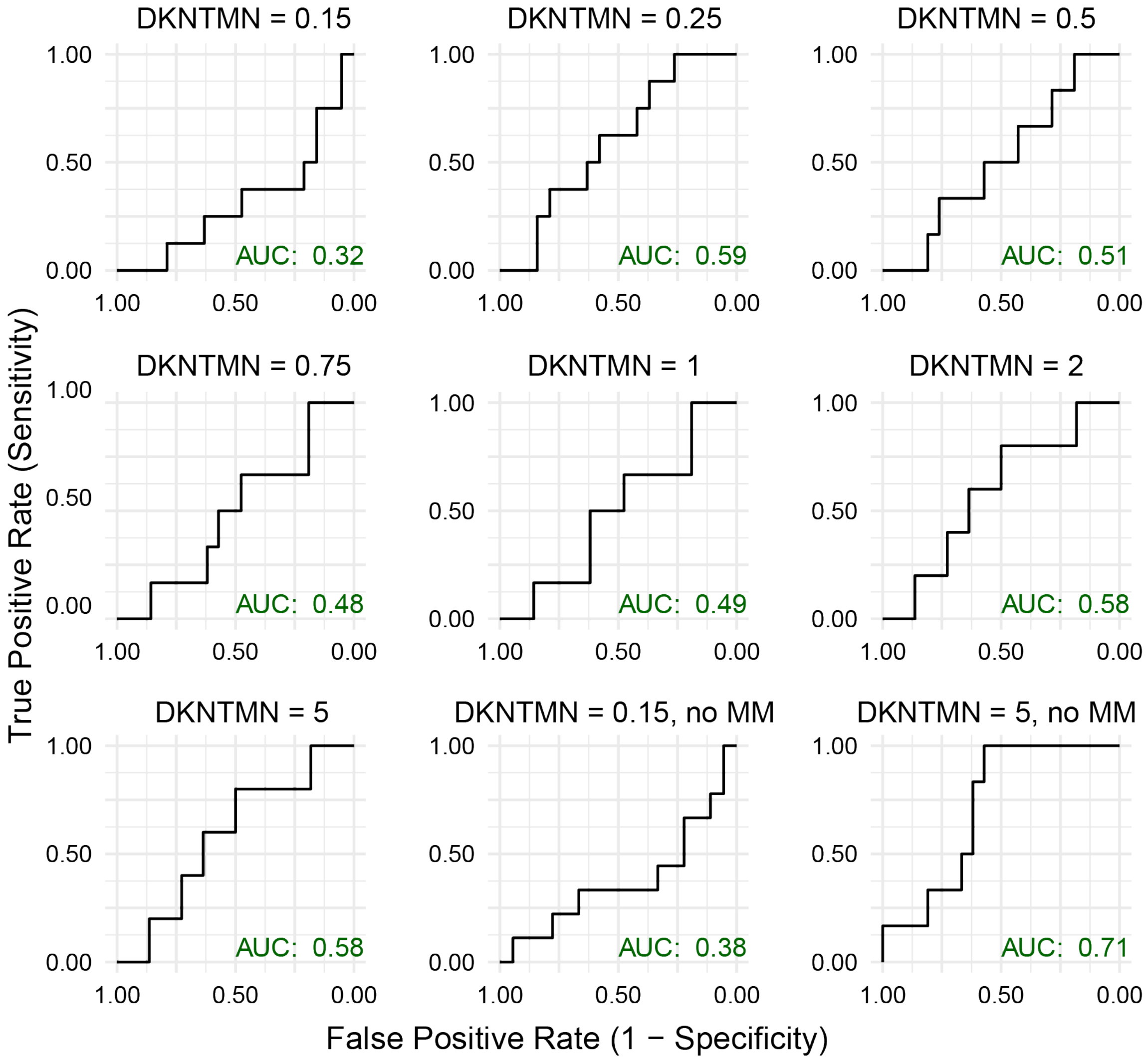


**Supp Figure 7.** Receiver operating characteristic (ROC) curves showing the performance of fitting quality number (FQN) in distinguishing false-positive 2HG detection for various spectral modeling approaches (i.e., various baseline stiffness defined using DKNTMN control parameter and with/without the inclusion of macromolecule, MM, signals in spectral fitting). Each plot includes the area under the curve (AUC). Only for the stiffest baseline modeling approach, excluding macromolecule signal models (DKNTMN = 5, no MM), FQN showed good performance (AUC = 0.71) in identifying false-positive 2HG detections in control spectra. The optimal cutoff for FQN was 1.4, yielding a sensitivity of 1 and a specificity of 0.57.

**MRSinMRS checklist**

The Minimum Reporting Standards for in vivo Magnetic Resonance Spectroscopy (MRSinMRS) checklist can be found in **Supplementary Table 1.**

**Supplementary Table 1**. MRSinMRS checklist for our multi-sequence MRS protocol.

| Site | | |
|---|---|---|
| 1. Hardware | | |
| a. Field strength [T] | 3 T | 3 T |
| b. Manufacturer | Philips | Philips |
| c. Model (software version if available) | Ingenia Elition (R 5.7.1) | Ingenia Elition (R 5.7.1) |
| d. RF coils: nuclei (transmit/ receive), number of channels, type, body part | 32-channel head coil | 32-channel head coil |
| e. Additional hardware | N/A | N/A |
| 2. Acquisition | | |
| a. Pulse sequence | $^1$H PRESS SVS | $^1$H PRESS SVS |
| b. Volume of Interest (VOI) locations | Patients: tumor and contralateral | Patients: tumor and contralateral |
| c. Nominal VOI size [$cm^3$, $mm^3$] | 8-27 ml | 8-27 ml |
| d. Repetition Time (TR), Echo Time (TE) [ms, s] | TR = 2000 ms, TE = 30 ms | TR = 2000 ms, TE = 97 ms |
| e. Total number of Excitations or acquisitions per spectrum<br>In time series for kinetic studies<br>i. Number of Averaged spectra (NA) per time-point<br>ii. Averaging method (e.g. block-wise or moving average)<br>iii. Total number of spectra (acquired / in time-series) | 128 | 256 |
| f. Additional sequence parameters<br>(spectral width in Hz, number of spectral points, frequency offsets)<br>If STEAM:, Mixing Time (TM)<br>If MRSI: 2D or 3D, FOV in all directions, matrix size, acceleration factors, sampling method | 2000 Hz, 2048 points | 2000 Hz, 2048 points<br>TE1/TE2 = 32/65 ms |
| g. Water Suppression Method | VAPOR | VAPOR |
| h. Shimming Method, reference peak, and thresholds for | $B_0$ map with shimmingtool[21] or vendor-provided automatic shimming | $B_0$ map with shimmingtool[21] or vendor-provided automatic shimming |

| | | |
|---|---|---|
| “acceptance of shim” chosen | (pencilbeam auto second order option) was used for shimming. | (pencilbeam auto second order option) was used for shimming. |
| i. Triggering or motion correction method (respiratory, peripheral, cardiac triggering, incl. device used and delays) | N/A | N/A |
| **3. Data analysis methods and outputs** | | |
| a. Analysis software | Osprey (v. 2.6.3), using the integrated LCModel fitting algorithm binary | Osprey (v. 2.6.3), using the integrated LCModel fitting algorithm binary |
| b. Processing steps deviating from quoted reference or product | Basis set created using MRSCloud, assuming ideal excitation, with sequence timings and refocusing RF pulse waveforms used in the actual sequence and effects of spatial localization considered (30 × 30 × 30 $mm^3$ volume, 41 x 41 points) | Basis set created using MRSCloud, assuming ideal excitation, with sequence timings and refocusing RF pulse waveforms used in the actual sequence and effects of spatial localization considered (30 × 30 × 30 $mm^3$ volume, 41 x 41 points) |
| c. Output measure (e.g. absolute concentration, institutional units, ratio)Processing steps deviating from quoted reference or product | Water referenced metabolite estimates (i.u.) | Water referenced metabolite estimates (i.u.) |
| d. Quantification references and assumptions, fitting model assumptions | The basis set included 2HG, alanine (Ala), ascorbate (Asc), aspartate (Asp), citrate (Cit), creatine (Cr), cystathionine (Cystat), ethanolamine (EA), gamma-aminobutyric acid (GABA), glycerophosphocholine (GPC), glutathione (GSH), glucose (Glc), glutamine (Gln), glutamate (Glu), glycine (Gly), myo-inositol (mI), lactate (Lac), N-acetylaspartate (NAA), N-acetylaspartylglutamate (NAAG), phosphocholine (PCh), phosphocreatine (PCr), phosphoethanolamine (PE), scyllo-inositol (sI), | The basis set included 2HG, alanine (Ala), ascorbate (Asc), aspartate (Asp), citrate (Cit), creatine (Cr), cystathionine (Cystat), ethanolamine (EA), gamma-aminobutyric acid (GABA), glycerophosphocholine (GPC), glutathione (GSH), glucose (Glc), glutamine (Gln), glutamate (Glu), glycine (Gly), myo-inositol (mI), lactate (Lac), N-acetylaspartate (NAA), N-acetylaspartylglutamate (NAAG), phosphocholine (PCh), phosphocreatine (PCr), phosphoethanolamine (PE), scyllo-inositol (sI), |

| | | |
|---|---|---|
| | serine (Ser), taurine (Tau). | serine (Ser), taurine (Tau). |
| 4. Data Quality | | |
| a. Reported variables (SNR, Linewidth (with reference peaks)) | All spectra are presented in Figure 1 and 5. | All spectra are presented in Figure 1 and 5. |
| b. Data exclusion criteria | existing artifacts, metabolite linewidth (FWHM) > 12 Hz | existing artifacts, metabolite linewidth (FWHM) > 12 Hz |
| c. Quality measures of postprocessing Model fitting (e.g. CRLB, goodness of fit, SD of residual) | There was no data rejected using a CRLB threshold. | There was no data rejected using a CRLB threshold. |
| d. Sample Spectrum | Figure 1 and 5 | Figure 1 and 5 |
